# COVID-19 Rapid Antigen Test at hospital admission associated to the knowledge of individual risk factors allow overcoming the difficulty of managing suspected patients in hospitals COVID-19 Rapid Antigen Test facilitates the management of suspected patients on hospital admission

**DOI:** 10.1101/2021.01.06.21249282

**Authors:** Priscilla S Filgueiras, Camila A Corsini, Nathalie BF Almeida, Jessica V Assis, Maria Luysa C Pedrosa, Alana K de Oliveira, Raquel NH Amorim, Daniel AP de Miranda, Lucélia A Coutinho, Sarah VC Gomes, Natália G Custódio, Douglas H da Silva, Gabriela PV Santos, Raphael A Silva, Maria Izabella VARC Medeiros, Priscila VCC Reis, Adelina J Lourenço, Cecília MF Bicalho, Raquel VR Vilela, Hércules P Neves, Gabriel R Fernandes, Rafaella FQ Grenfell

**Author notes:** Corresponding author. 1715 Avenida Augusto de Lima, Belo Horizonte, Minas Gerais, Brazil. 30.190- 002.

## Abstract

Early diagnosis of SARS-CoV-2 is essential to limiting the spread of the virus and managing infected patients during hospitalization. The sensitivity of RT-qPCR is contested by the fact that it is time-consuming, executed by trained technicians in proper environment for material extraction. Here, we evaluated the first SARS-CoV-2 antigen test recommended by the World Health Organization at September, 2020 as an alternative for immediate diagnosis of symptomatic and suspected patients at a hospital in Brazil during the epidemic peak. All patients were submitted to RT-qPCR and rapid antigen test using nasopharyngeal swabs rigorously collected at the same time. Demographics, baseline comorbidities, symptoms and outcomes were considered. Prediction analysis revealed that previous stroke, chronic obstructive pulmonary disease, desaturation and tachypnea were the most relevant determinants of the death of COVID-19 patients. Comparison between the rapid antigen test and RT-qPCR revealed an overall PPV of 97%, extended to 100% when performed between 4 and 15 days of symptoms, with an accuracy of 90-91% from days 1 to 7 and a ‘Substantial’ agreement. The rapid antigen test presented no inconclusive result. Among the discordant results and RT-qPCR inconclusives, 72% presented bilateral multifocal ground-glass opacities on imaging and other exams alterations. The median time to obtain RT-qPCR results was 83.6 hours, against 15 minutes for the rapid test, precious time for deciding on patient isolation and management. Knowledge of the risk factors and a rapid diagnosis upon patient admission is critical to reduce mortality of COVID-19 patients, hospital internal costs and in-hospital transmission.

## Introduction

Multiple pneumonia cases of unknown origin were described in December 2019 in China, which some days later were related to a novel coronavirus. The severe acute respiratory syndrome caused by the new virus (SARS-CoV-2) was then denominated coronavirus disease 2019 (COVID-19) (1). Since its first notification, the virus has spread around the world in record time despite rigorous attempts to contain the disease. In March 8, 2020, statistical data showed that the outbreak of COVID-19 constitutes an epidemic threat worldwide, as declared by the World Health Organization (WHO) (2).

Real-time quantitative reverse-transcriptase polymerase chain reaction (RT-qPCR) assay is the gold standard in the diagnosis of SARS-CoV-2 infection (3-5) and it is an indicator of isolation, discharge, or transferring for patients diagnosed with COVID-19 (6). However, the detection of the viral nucleic acid takes hours to be completed and requires specialized instrument and expertise. In addition, diagnostic reports may take days to reach the patient and medical team, weakening decision making about hospitalization and isolation of patients, which has great impact on hospital’s total operating costs. While waiting for the diagnosis of patients with suspected COVID-19, hospital are forced to decide for isolation which, in turn, involves approximately one-third of hospitals’ total operating costs and several areas as emergency room, intensive care unit, outpatient clinic, laboratory, diagnostic radiology, and general routine inpatient care (7).

For COVID-19 positive individuals, the age, the existence of a previous stroke, and chronic obstructive pulmonary disease were the most important comorbidities associated with the death event. Among the symptoms, desaturation and tachypnea were the most relevant determinants of the death of COVID-19 patients. These markers were more important to predict the outcome of COVID-19 positive patients than COVID-19 negative patients

The introduction of rapid antigen tests in hospitals and clinical laboratories is recent, as is the assessment of its accuracy. In the interim guidance of September 11, 2020, WHO has presented this option as a new technology for COVID-19 detection that is simpler and faster to perform than currently-recommended nucleic acid amplification tests (8). Rapid antigen tests are based on the direct detection of SARS-CoV-2 viral proteins in nasopharynge, oropharynge or possibly nasal swabs. Platforms are centered on a lateral flow immunoassay that provides results in up to 30 minutes. Though the sensitivity of these antigen detection tests are under evaluation in some countries, they offer the possibility of rapid and early detection of the most infectious COVID-19 cases with no requirement of special equipment or expertise, enabling conscious decision about isolation of patients in appropriate settings.

Considering that highly sensitive and specific tests are crucial to identify and manage COVID-19 patients and implement control measures to limit the outbreak and the appearance of new epidemic peaks, this work describes the performance of SARS-CoV-2 rapid antigen test (ECODiagnostica, Brazil) when introduced in a COVID-19 specialized hospital in Brazil. The selected test combines immunochromatography with monoclonal anti-SARS-CoV-2 antibodies conjugated with gold particles forming an antigen-antibody complex when detecting the viral nucleocapsid (N) viral protein. Performance of the test was correlated to RT-qPCR results, demographics, baseline comorbidities, baseline symptoms, and outcomes of the first sequentially hospitalized and tested patients with suspected COVID-19 in a hospital that provides health services for the Unified Health System (*Sistema Único de Saúde* - SUS) of the Brazilian Federal Government.

## Materials and methods

### Patients and sample collection

Patients with clinical features and suspected COVID-19 were enrolled in this study after being attended by emergency room clinicians of a reference hospital in Belo Horizonte, Brazil during the epidemic peak in Minas Gerais state. All patients were examined for SARS-CoV-2 by RT-qPCR and SARS-CoV-2 rapid antigen test using nasopharyngeal swabs rigorously collected at the same time of the first days of symptoms onset considering the WHO recommendations (9). One sample of each patient was submitted to RT-qPCR diagnosis conducted by the Minas Gerais state referenced laboratories (Belo Horizonte, Brazil), while second sample was immediately tested by the rapid antigen test. Individual patient information was collected from the hospital electronic medical records and included demographic aspects, primary symptoms, comorbidities, exams during hospitalization, RT-qPCR result and outcomes.

### Detection of SARS-CoV-2 viral antigen by a rapid antigen test

The SARS-CoV-2 rapid antigen test used in this work was the first method approved by the National Health Surveillance Agency of the Brazilian Federal Government, COVID-19 Ag ECO Test (ECODiagnostica, Brazil). The nasopharynge sample for analysis was obtained by dipping a swab through the nostril parallel to the palate. Swab was gently rubbed and rolled, and was slowly removed and introduced into the extraction buffer provided by the kit, where swab was rotated at least 10 times. Three drops of the mixture were added to the sample port of the antigen assay and the results were interpreted after a 15-min incubation.

### RT-qPCR for SARS-CoV-2 virus

The RT-qPCR assay used was based on the detection of nucleic acid from SARS-CoV-2 in upper respiratory samples collected with a nasopharyngeal swab as previously described using the Allplex™ 2019-nCov Assay kit (Seegene Inc., Republic of Korea) and was performed by the Minas Gerais Reference Center for the Diagnosis of COVID-19 (10). The detected viral targets include Gene E, Gene RdRP and Gene N. MS2 bacteriophage was used as an internal control. Pre-processed samples were stored under refrigeration until RNA extraction with magnetic beads. Each 25 μL reaction mixture contained 8 μL of extracted RNA, 5 μL of 2019-nCoV MOM oligonucleotides, 5 μL of nuclease-free water, 5 μL of Real-time One-step Buffer (5X) and 2 μL of Real-time One-step Enzyme (Thermo Fisher Scientific, USA). The reading was performed on a 7500 Fast Real-Time PCR System (Thermo Fisher Scientific, USA), where readings were defined as: Gene E - FAM, Gene RdRP - Cal Red 610, Gene N - Quasar 670 and IC - HEX. Cycles were defined as: 1 cycle of 50°C for 20 min, 1 cycle of 95°C for 15 min, 45 cycles of 94°C for 15 sec and 58°C for 30 sec. Amplifications were analyzed using Seegene Launcher software (Seegene Inc., Republic of Korea).

### Ethical statement

Written informed consent was obtained from each enrolled patient at the hospital. This study was reviewed and approved by the Ethical in Human Research Committee of Oswaldo Cruz Foundation and the National Brazilian Ethical Board (CONEP N. 30428720.3.0000.5091).

### Data analyses

The data retrieved from the medical records were explored using the Factor Analysis for Mixed Data (FAMD) from the FactoMineR package (11). The comorbidities and symptoms were used as variables to build models to predict the outcome - death or hospital discharge - independently for COVID-19 positive and COVID-19 negative patients. The Random Forest algorithm implemented in R (12) was used to create the models and measure de predictors’ importance. Eighty per cent of the patients were randomly selected as the training set, and the algorithm performed 20 repetitions of cross-validation. The predictors’ importance was calculated by the Mean Decrease in Gini. The procedure was repeated 100 times to assess the Mean Decrease variation in different training sets. We compared the variables’ importance between the models built only with COVID-19 positive patients and built only with COVID-19 negative patients. The t-test (p < 0.05) pointed out the predictors that are important to predict the outcome for COVID-19 positive patients. Differences on mortality rates and data of laboratory tests were calculated, respectively, by student *t* test and Fisher’s F-Test for two variances (p ≤ 0.05) on Minitab Software. Sensitivity, specificity, accuracy, positive and negative predictive values were calculated by MedCalc statistical software (p ≤ 0.05) (13). Charlson Comorbidity Index (14) was estimated by MdCalc online calculator (15). The agreement between methods was assessed by Kappa Index calculated by GraphPad (GraphPad Software, Inc., USA): k < 0.01 no agreement; k = 0.01-0.20 ‘poor’; k = 0.21-0.40 ‘fair’; k = 0.41-0.60 ‘moderate’; k = 0.61-0.80 ‘substantial’; k = 0.81-1.00 ‘almost perfect’ (16).

## Results

### Patient characteristics and clinical outcomes

A total of 150 hospitalized patients (57 male, 7-89 years; 93 female, 16-99 years) with suspected COVID-19 were identified between June to August, 2020. From whom 55 (36.7%) patients were confirmed with COVID-19 with positive RT-qPCR test result. Of all COVID-19 positive patients, the median age was 62 years (ranging from 29 to 91 years), 22 (40.0%) were male, 33 (60.0%) were female and 36 (65.5%) were critically ill. The median time to obtain polymerase chain reaction testing results was 83.6 hours (ranging from 24.2 to 182.3 hours). The most common comorbidities were hypertension (71, 47.3%), diabetes (36, 24.0%), cancer (32, 21.3%), chronic kidney disease (20, 13.3%), heart disease (11, 7.3%) and chronic obstructive pulmonary disease (11, 7.3%). Twenty seven (18.0%) of the hospitalized patients did not present any comorbidity. The median score on the Charlson Comorbidity Index (11) was 17 points, which corresponds to a 58.6% estimated 10-year survival and reflects a significant comorbidity burden for these patients. At triage, 72 patients (48.0%) presented dyspnea, 52 (34.7%) dry cough, 50 (33.3%) fever, 49 (32.7%) myalgia, 25 (16.7%) asthenia and 24 (16.0%) productive cough, as presented in Table 1. One hundred and twenty seven (84.7%) patients had more than 2 symptoms associated, while 15 (10%) patients had between 5 and 7 associated symptoms.

**Table 1.**
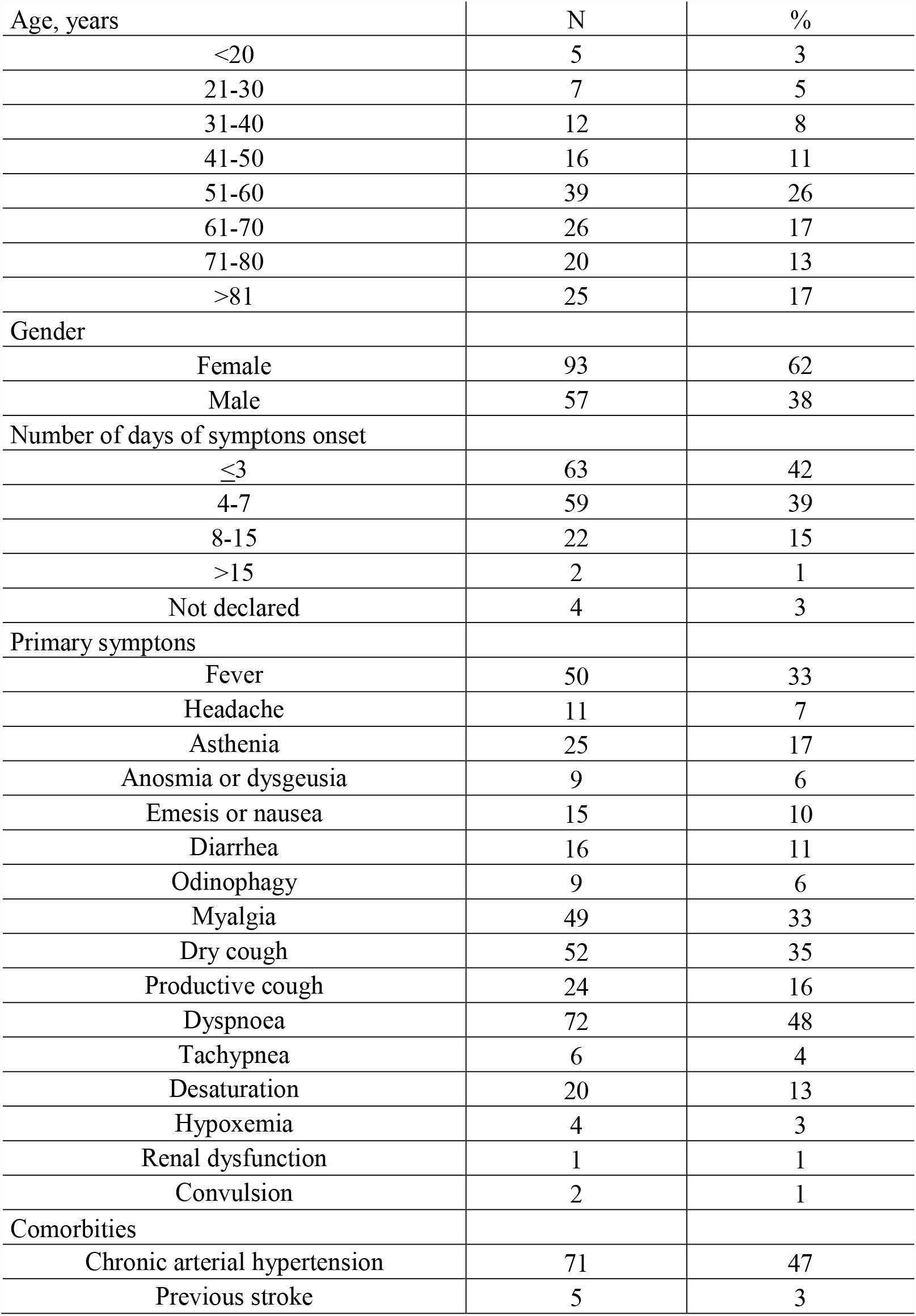
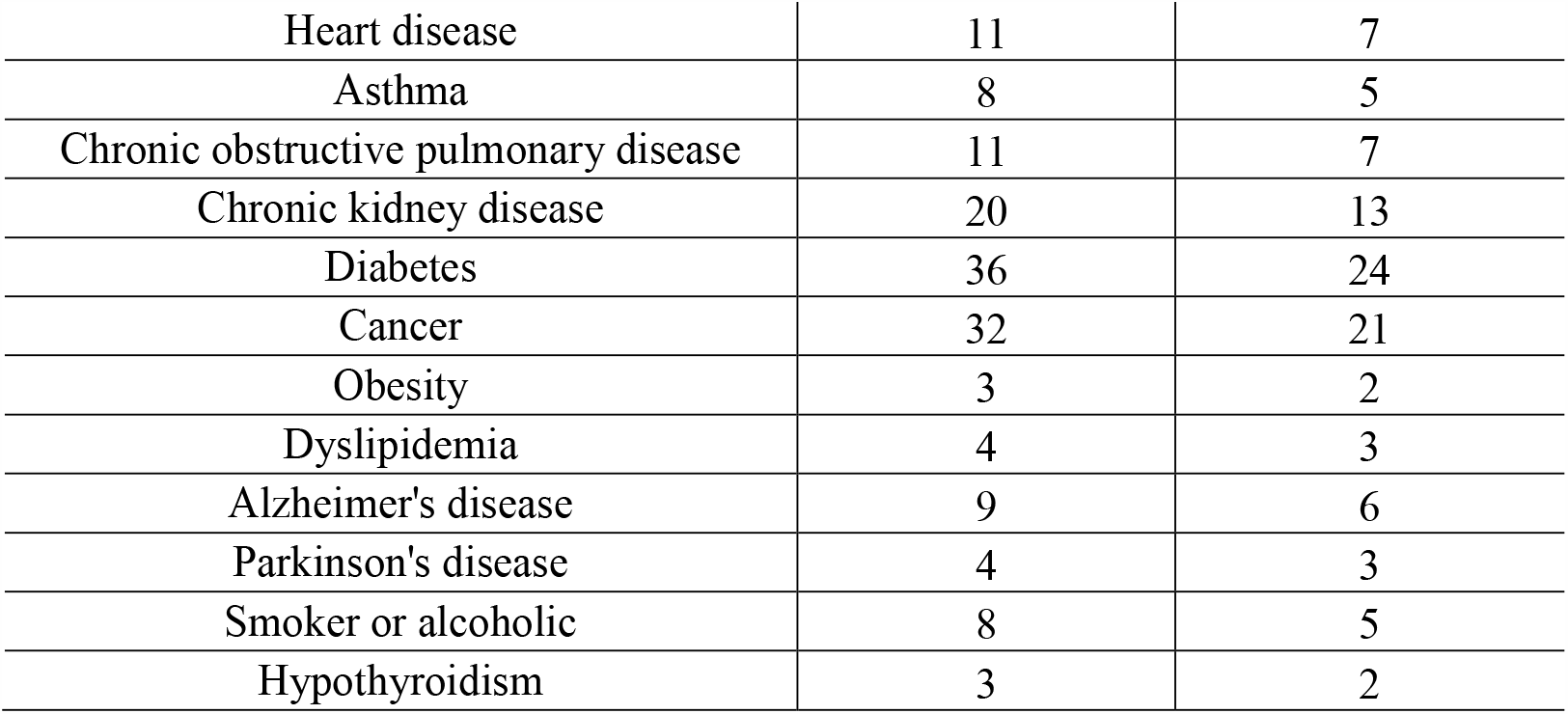
Baseline characteristics of patients admitted to a hospital in Brazil with suspect COVID-19

Length of stay for those COVID-19 positive who died and were discharged alive is included in Table 2. Two patients remained hospitalized at the final study follow-up date, a female positive for COVID-19 hospitalized in isolation and a male negative for COVID-19 hospitalized in ICU. The median follow-up at time of censoring was 13 days. Mortality was 0% for male patients younger than 40 years and female younger than 50 years and between 61-70 years, 1.8% (1/55) for male patients between 41-50, 51-60 years and older than 80 years and also for female patients between 51-60 years old when all hospitalized patients were grouped. Male between 61-70 and 71-80 years and female between 71-80 and older than 81 years reached mortality of 3.6% (2/55). The case fatality rate was 21.8% (12/55) for COVID-19, and the recovery rate was 78.2% (43/55). Mortality rates were equal for male and female patients when considering all age groups (p = 0.641). Length of hospitalization mean was 8.7 (SD = 7.8) and 7.7 (SD = 7.1) days respectively for patients positive and negative for COVID-19.

**Table 2.** Outcome and length of stay of the 55 patients hospitalized with COVID-19

|  | Patients dead or discharged alive |  |  |  |  |  |
| --- | --- | --- | --- | --- | --- | --- |
|  | Died, N (%) |  | Length of stay<br>among those<br>who died, mean<br>(SD)* | Discharged alive, N (%) |  | Length of stay<br>among those who<br>discharged alive,<br>mean (SD)* |
|  | Male | Female |  | Male | Female |  |
| Age intervals |  |  |  |  |  |  |
| <20 | 0 (0%) | 0 (0%) | NA | 0 (0%) | 0 (0%) | NA |
| 21-30 | 0 (0%) | 0 (0%) | NA | 0 (0%) | 1 (1.8%) | 1.0 (0.0) |
| 31-40 | 0 (0%) | 0 (0%) | NA | 4 (7.3%) | 1 (1.8%) | 6.9 (3.1) |
| 41-50 | 1 (1.8%) | 0 (0%) | 19.0 (0.0) | 2 (3.6%) | 3 (5.5%) | 6.0 (2.6) |
| 51-60 | 1 (1.8%) | 1 (1.8%) | 5.0 (0.0) | 4 (7.3%) | 13 (23.6%) | 13.8 (6.2) |
| 61-70 | 2 (3.6%) | 0 (0%) | NA | 1 (1.8%) | 3 (5.5%) | 26.4 (9.4) |
| 71-80 | 2 (3.6%) | 2 (3.6%) | 20.5 (10.2) | 2 (3.6%) | 4 (7.3%) | 24.6 (12.5) |
| >81 | 1 (1.8%) | 2 (3.6%) | 23.2 (10.2) | 3 (5.5%) | 2 (3.6%) | 9.8 (3.6) |
| Total | 7 (12.7%) | 5 (9.1%) |  | 16 (29.1%) | 27 (49.1%) |  |

\* Length of stay begins with admission day and ends with death or discharge date.

At the hospitalization date, 116 patients were submitted to lung x-ray and chest tomography scans. According to the CT images, all COVID-19 positive individuals that demonstrated an alteration on imaging presented bilateral pneumonia (19/19) with typical findings of patchy ground-glass shadows in lung, while 5/11 (45.5%) of COVID-19 negative individuals presented same alteration. As shown in Table 3, data of laboratory tests revealed no difference between COVID-19 positive and negative patients on < 60 or ≥ 60 years old groups on any parameter evaluated. Data from blood routine showed that all patients with alterations on hemoglobin and lymphocytes presented values below the normal range. On the other hand, leukocytes and neutrophils were elevated in all cases. Platelet was suppressed in 19/64 (29.7%) and 8/47 (17.0%), and elevated in 6/64 (9.4%) and 1/47 (2.1%) cases, respectively for COVID-19 negative and COVID-19 positive groups. Elevated prothrombin time was identified only on 5/28 (17.9%) of COVID-19 negative patients under 60 years, while 8/26 (30.8%) of same age COVID-19 positive group. There were no significant differences in lactate, D-dimer, T and I troponin, AST and ALT in the groups. Creatinine and urea were elevated in a minority of COVID-19 negative and positive patients and presented no significant difference when ages were compared. Bilirubin was not altered in any of the individuals, with only one COVID-19 negative patient presenting low bilirubin value (Table 3).

**Table 3.**
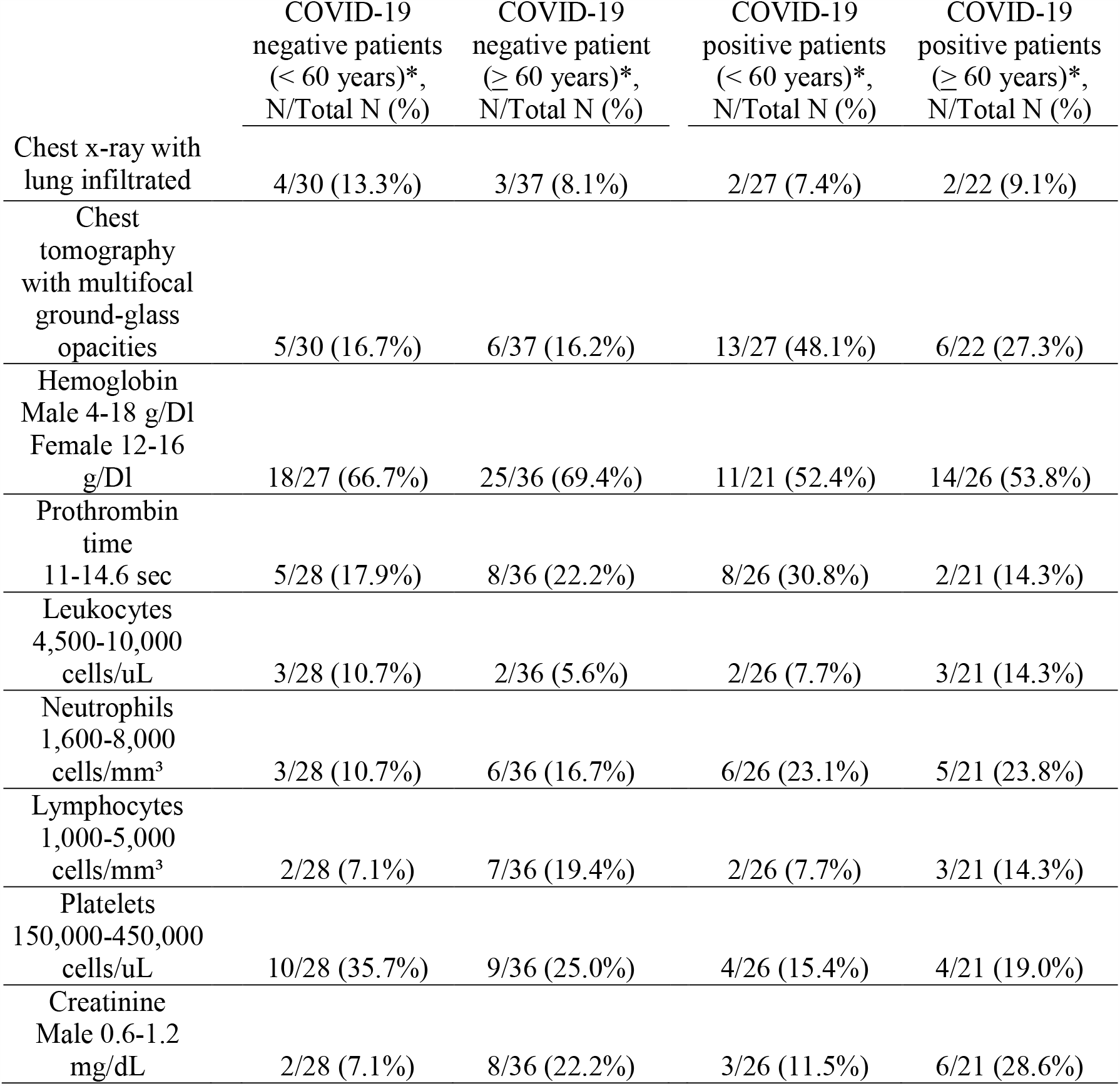
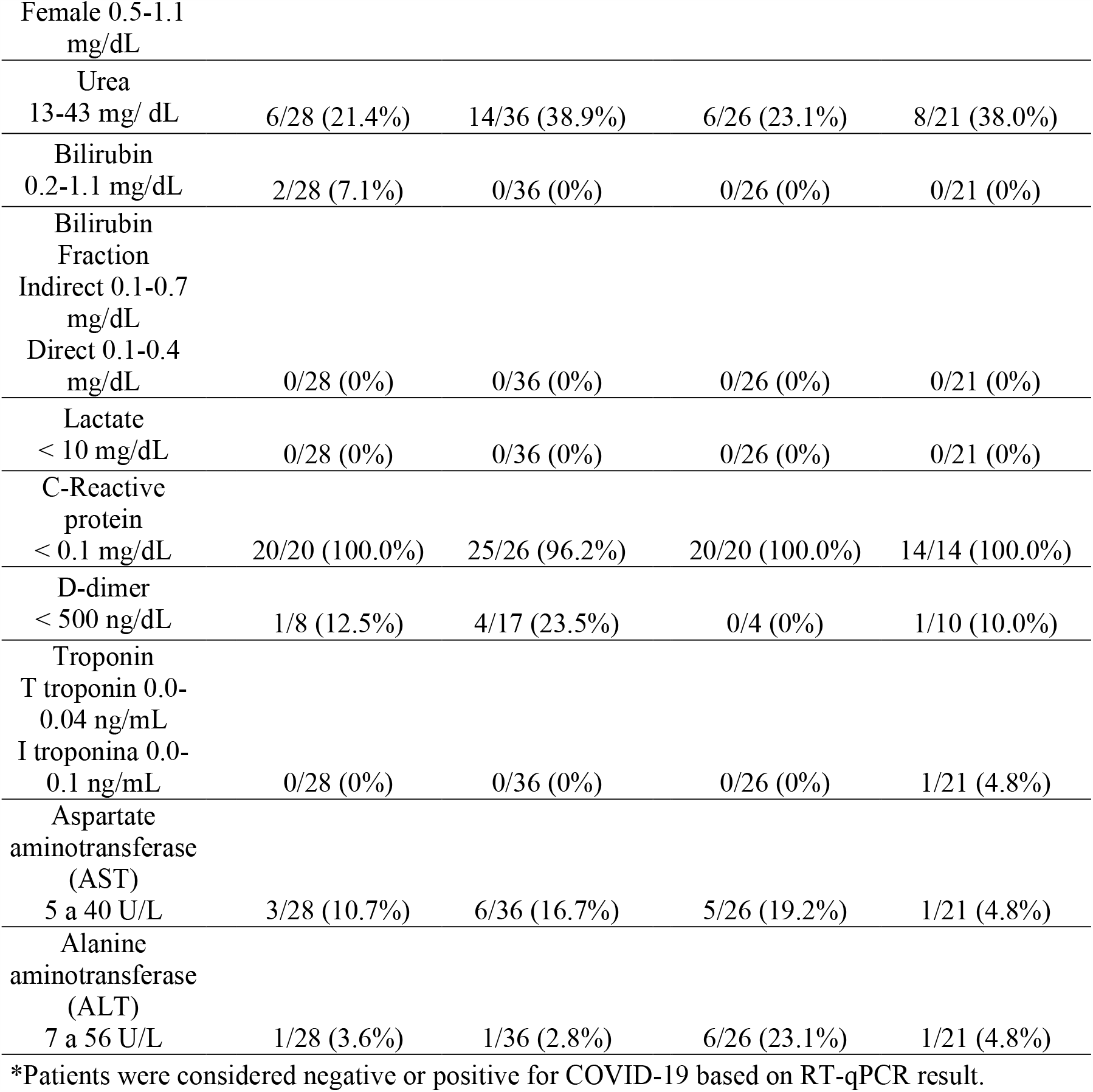
Number and percentage of patients negative or positive for COVID-19 with values that exceeded the standard limit range on laboratory tests results

### Factor analysis of Mixed Data - Comorbidities

In order to identify patterns in the distribution of comorbidities and symptoms across COVID-19 patients, we performed a Factor Analysis. The EigenValue, which measures the variation along the axis, could explain 14.7% and 11.0% of the variation in axis 1 and 2, respectively. The ordination plot shows the patients colored by age, and the shape represents the patient’s outcome - death or hospital discharge (Figure 1A). The boxplots report the patients’ distribution along the axis (Figure 1A). The patients who died are concentrated at the positive portion of the axis 1 while equally distributed along the axis 2. We also observe the distribution of ages along the axis 1, with the older patients concentrated on the positive side of the axis 1. Each comorbidity’s contribution was calculated, and the most important variables were plotted in the ordination plot. The bar plots show the most critical drivers of the axis 1 and 2 (Figure 1B). The first dimension was mainly driven by age. Chronic arterial hypertension, diabetes, and heart disease were essential contributors to guide individuals to the positive side of the axis 1. The most important drivers of axis 2 are dyslipidemia, cancer, and Alzheimer’s disease.

**Figure 1.**
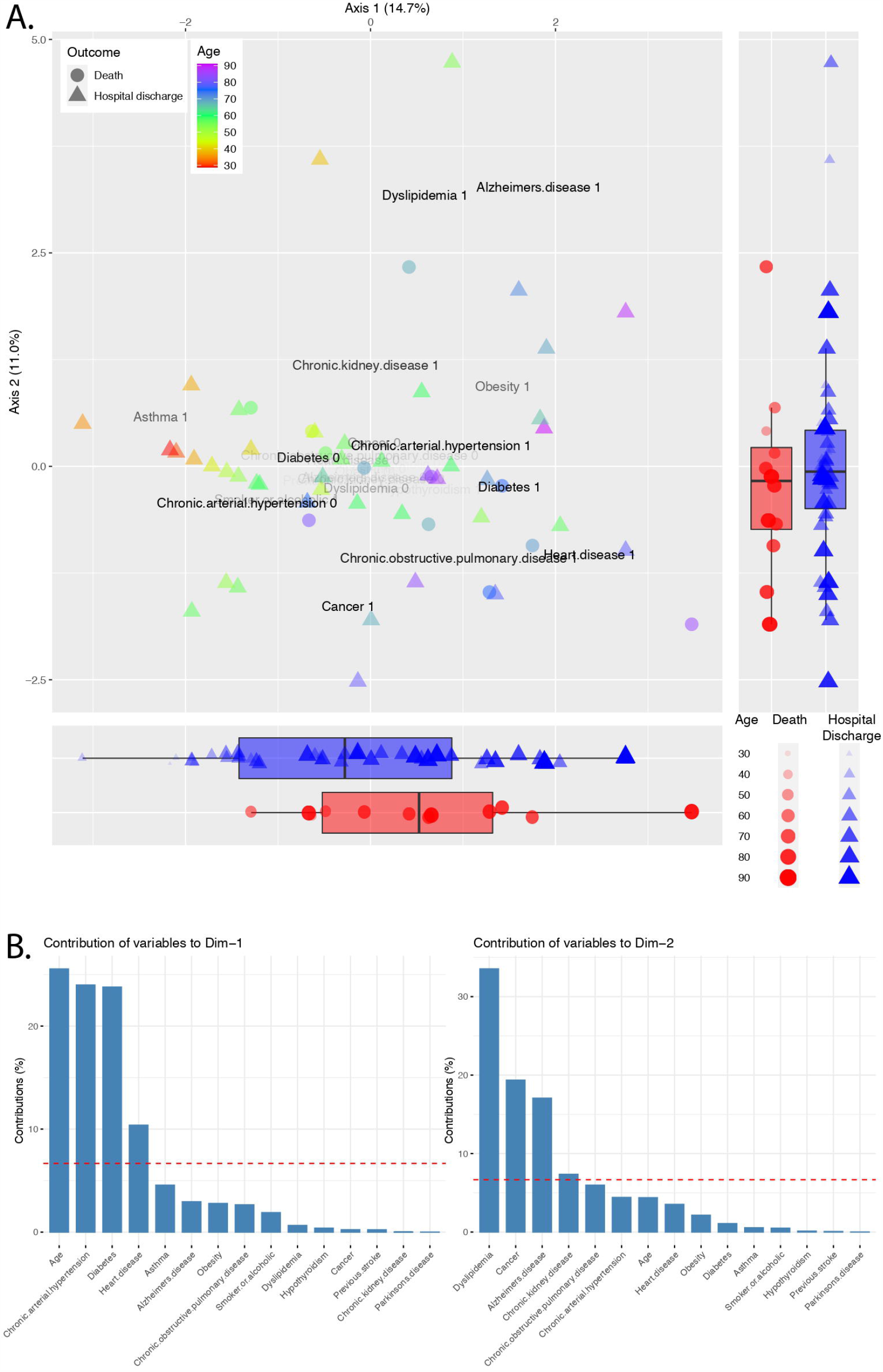
Factor analysis of mixed data of comorbidities across COVID-19 patients. A. Ordination plot shows patients colored by age and the patient’s outcome - death or hospital discharge – represented by shapes. The opacity of the variable name shows the relative contribution for the individual dispersion. Boxplots report patients’ distribution along the axis. B. The bar plots pictures the contribution of each variable to the variance across axis 1 and 2. The red dashed line represents the expected contribution if each variable contributes equally.

### Factor analysis of Mixed Data - Symptoms

A similar analysis was used to identify the patterns and factors related to patients’ symptoms (Figure 2). The Eigenvalue for the first two dimensions were 15.3% and 12.4% for axis 1 and axis 2, respectively. The boxplots reflect a concentration of deaths on the positive section of the axis 2and the axis 1 negative side (Figure 2A). Again, we observe the distribution of the ages along the axis 1, with a concentration of older patients on the left side of the plot. The variables that most contributed to driving the individuals along the axis 1 are productive cough, odinophagy, headache, myalgia, nausea, age, and anosmia; while through the axis 2 were dry cough, odinophagy, dyspnoea, myalgia, asthenia, tachypnea, age, productive cough, and nausea (Figure 2B).

**Figure 2.**
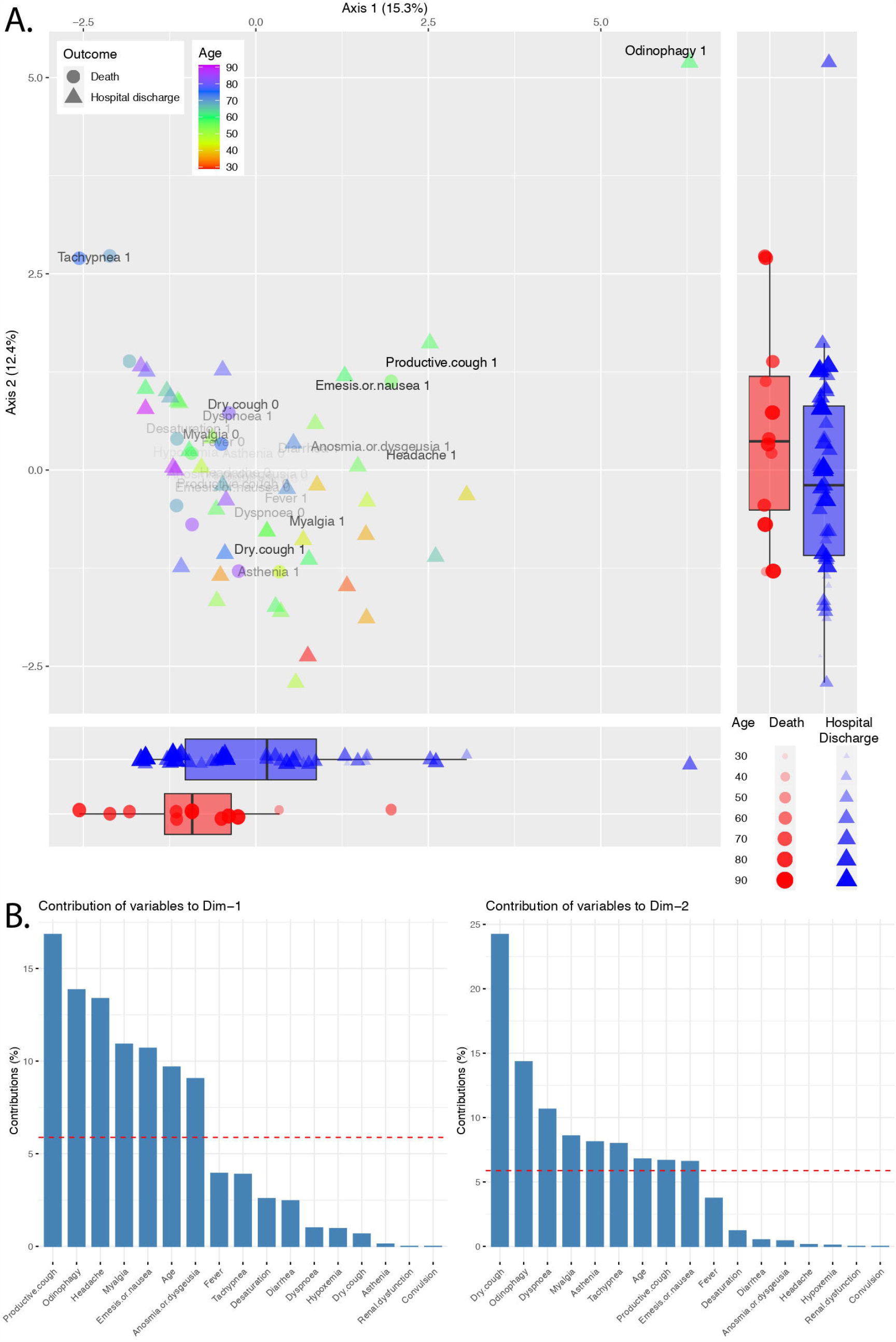
Factor analysis of mixed data of symptoms across COVID-19 patients. A. Ordination plot shows patients colored by age and the patient’s outcome - death or hospital discharge – represented by shapes. The opacity of the variable name shows the relative contribution for the individual dispersion. Boxplots report patients’ distribution along the axis. B. The bar plots pictures the contribution of each symptom to the variance across axis 1 and 2. The red dashed line represents the expected contribution if each variable contributes equally.

### Predictive variables for patients’ outcome

In order to identify variables with predictive potential, we built random forest models to predict the outcome - death or hospital discharge - for COVID-19 positive and negative patients. The mean Gini Decrease was calculated to quantify the importance of each variable. For COVID-19 positive individuals, the age, the existence of a previous stroke, and chronic obstructive pulmonary disease were the most important comorbidities associated with the death event (Figure 3A). Among the symptoms, desaturation and tachypnea were the most relevant determinants of the death of COVID-19 patients (Figure 3B). These markers were more important to predict the outcome of COVID-19 positive patients than COVID-19 negative patients (p < 0.05).

**Figure 3.**
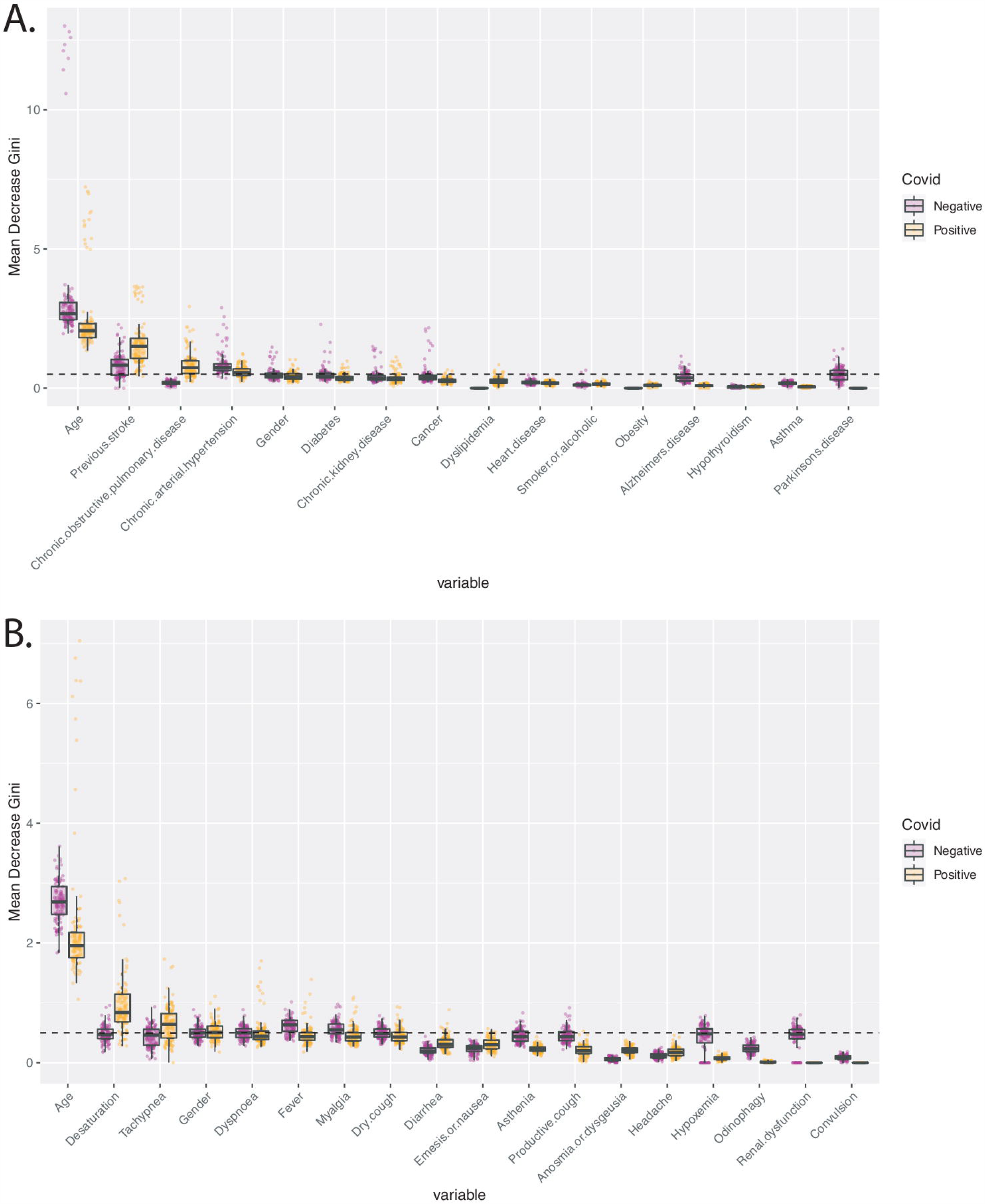
Mean Decrease Gini values for prediction of the outcome - death or hospital discharge - for COVID-19 positive and negative hospitalized patients. A. Boxplots picture the importance of comorbidities in predicting the outcome of Covid-19 negative – purple points -, and positive patients – orange points. The dashed line marks a Mean Decrease Gini value of 0.5. B. Boxplots picture the importance of symptoms in predicting the outcome of Covid-19 negative – purple points -, and positive patients – orange points. The dashed line marks a Mean Decrease Gini value of 0.5.

### Performance of Ag test in diagnosing COVID-19

We collected two nasopharyngeal samples from each of the 150 patients at the same time. The first sample was immediately submitted to SARS-CoV-2 rapid antigen test and the second sample was directed to RT-qPCR. According to RT-qPCR results, 55 samples were positive and 84 were negative, while 11 samples presented inconclusive results. Cycle threshold (Ct) values were only made available by the Reference Center for 5 patients thus it was not possible to determine the median ct value. SARS-CoV-2 rapid antigen test revealed 40 positive samples and 110 negative samples. Diagnostic results and agreement/disagreement between methods are described on Table 4 and Table 5. Amongst the 139 patients with conclusive results for RT-qPCR, 38/55 (69.1%) were both positive while 83/84 (98.8%) were both negative, 17/55 (30.9%) had a positive RT-qPCR result only and 1/84 (1.2%) of them was positive only for the rapid test. Sensitivity and specificity of SARS-CoV-2 rapid antigen test was 69% and 98%, respectively. The probability that the disease was present when the test was positive and not present when the test was negative was determined by predictive values (97% and 83%, respectively), while accuracy was 87%. The performance of SARS-CoV-2 rapid antigen test in diagnosing COVID-19 in different age and gender groups is reported in Table 4. No significant difference was seen on the test performance when evaluating patients by age (group 1: 60-99 years, group 2: 7-59 years) with accuracies of 89% and 86%, respectively. Sensitivity (79%), positive and negative predictive values (100% and 88%, respectively) and accuracy (92%) of the rapid test in diagnosing COVID-19 were significantly higher in female patients (p = 0.001). Together, number of days with symptoms was decisive on the test performance. Accuracies of 91% and 90% were reached when diagnosis was done on days 1-3 and 4-7 of symptoms in that order, while only 67% when diagnosis were performed after 8 days of symptomatology (p = 0.001). Sensitivity and negative predictive value were significantly low when diagnosis were performed after 8 days of symptoms onset (p = 0.001).

**Table 4.** Performance of SARS-CoV-2 rapid antigen test in the diagnosis of COVID-19 in comparison to RT-qPCR as the reference standard.

| Parameter | Results |  |  |  | Test Performance, % (95% CI) |  |  |  |  |  |
| --- | --- | --- | --- | --- | --- | --- | --- | --- | --- | --- |
|  | TP | TN | FP | FN | Sensitivity | Specificity | PPV | NPV | Accuracy | Kappa Index |
| Overall | 38 | 83 | 1 | 17 | 69 (55-81) | 98 (94-100) | 97 (84-100) | 83 (76-88) | 87 (80-92) | 0.72 (0.60-0.84) |
| Age |  |  |  |  |  |  |  |  |  |  |
| ≥ 60 years | 16 | 46 | 0 | 8 | 67 (45-84) | 100 (92-100) | 100 | 85 (77-91) | 89 (79-95) | 0.72 (0.55-0.90) |
| < 60 years | 22 | 37 | 1 | 9 | 71 (52-86) | 97 (86-100) | 96 (76-99) | 80 (70-88) | 86 (75-93) | 0.70 (0.53-0.87) |
| Gender |  |  |  |  |  |  |  |  |  |  |
| Male | 12 | 30 | 1 | 10 | 55 (32-76) | 97 (83-100) | 92 (63-99) | 75 (65-83) | 79 (66-89) | 0.55 (0.33-0.77) |
| Female | 26 | 53 | 0 | 7 | 79 (61-91) | 100 (93-100) | 100 | 88 (80-94) | 92 (84-97) | 0.82(0.70-0.95) |
| N of days with symptoms |  |  |  |  |  |  |  |  |  |  |
| <3 | 9 | 44 | 1 | 4 | 69 (39-91) | 98 (88-100) | 90 (56-99) | 92 (83-96) | 91 (81-97) | 0.73 (0.51-0.95) |
| 4-7 | 24 | 27 | 0 | 6 | 80 (61-92) | 100 (87-100) | 100 | 82 (69-90) | 90 (79-96) | 0.79 (0.64-0.95) |
| 8-15 | 4 | 10 | 0 | 7 | 36 (11-69) | 100 (69-100) | 100 | 59 (48-69) | 67 (43-85) | 0.35 (0.05-0.65) |
| >15 | 0 | 1 | 0 | 0 | - | 100 (3-100) | - | 100 | - | - |

TP = true positive, TN = true negative, FP = false positive, FN = false negative, CI = confidence intervals, PPV = positive predictive value, NPV = negative predictive value.

**Table 5.** Reproducibility and disagreement of SARS-CoV-2 rapid antigen test and RT-qPCR

|  | N/Total N (%) |
| --- | --- |
| Agreement | 121/139 (87.1) |
| Disagreement | 18/139 (12.9) |
| RT-qPCR positive + antigen test negative | 17/139 (12.2) |
| RT-qPCR negative + antigen test positive | 1/139 (0.7) |
| Inconclusive results | 11/150 (7.3) |
| RT-qPCR | 11/150 (7.3) |
| SARS-CoV-2 rapid antigen test | 0/150 (0) |

Agreement between RT-qPCR and SARS-CoV-2 rapid antigen test was substantial with a kappa index of 0.72. There was no significant difference in reproducibility of tests by age, but once more a moderate agreement was found for male patients (κ = 0.55) while an almost perfect agreement was found for female patients (κ = 0.82). Differences were observed between RT-qPCR and SARS-CoV-2 rapid antigen test when considering the time of detection with a substantial agreement when diagnosis were made from day 1 to 7 of symptoms onset (κ = 0.73 and 0.79, respectively for <3 and 4-7 days) and a poor agreement for diagnosis performed after day 8 (κ = 0.35).

Discordant results among tests were observed for 18/139 samples (12.9%). Amongst the 17 patients positive only for RT-qPCR (5 male, 4 female, <60 years; 5 male, 3 female, ≥60 years), 1 was diagnosed on day 2 after symptoms onset, 3 on day 3, 6 on days 4-5, 6 on days 8-10 and 1 on day 15. Three of these patients died after 3-24 days of hospitalization (2 male, 1 female, 69-85 years), with no lung infiltrate on chest x-ray or multifocal ground-glass opacities on chest tomography and, no alteration on other exams, except for an elevated urea and c-reactive protein on one patient. There were 7/14 patients with hospital discharge that presented positive chest tomography findings (5 male, 4 female, <60 years; 3 male, 2 female, ≥60 years) but no lung infiltrate on chest x-ray, and only 4 presented other exams alterations including elevated c-reactive protein and/or prothrombin time. From the other 7 patients with hospital discharge presenting no alterations on imaging, one presented no other alteration on any exam while 6 showed elevated prothrombin time, elevated neutrophils, decreased lymphocytes, elevated c-reactive protein and/or elevated transaminases. From all the discordant results, none of the patients presented low saturation or needed ventilation or oxygen supply.

One RT-qPCR negative patient tested positive for SARS-CoV-2 rapid antigen test after 2 days of symptoms. This patient were admitted with fever, dry cough and dyspnea, presented chest tomography with multifocal ground-glass opacities on both sides and reduced hemoglobin. Despite dyspnea, he did not required oxygen supply and remained hospitalized for 5 days before hospital discharge.

From the 11 RT-qPCR inconclusive results, 5 were on the first and second days of symptomatology, 1 of them were on the 8th and 1 on the 15^th^ of symptoms. Only 2 patients were on the 5^th^ day of symptoms while no report was found for 2 of them. Two patients presented chest tomography with bilateral multifocal ground-glass opacities and 4 needed oxygen supply due to strong dyspnea and desaturation (2 of them evolved to death). Only 1/11 of these patients with inconclusive RT-qPCR were positive for SARS-CoV-2 rapid antigen test. This patient (1 day of symptoms) presented odynophagia and dry cough.

## Discussion

In the ongoing pandemic context of COVID-19, diagnostic testing for SARS-CoV-2 is critical for limiting the spread of the virus as well as managing infected patients during hospitalization. In the first months of the pandemic, there was considerable challenge regarding the use of nucleic acid test or clinical characteristics as reference standards to make a definitive diagnose of patients (17). Many advances have been made and RT-qPCR has become a reliable method of detection when performed on days 3 to 8 of symptoms onset. Despite its sensitivity, RT-qPCR is a time-consuming method limited by several practical issues, including the need for specialized operators and certified laboratories, making its use particularly challenging in resource-limited settings. This information is particularly relevant to the reality of developing countries that face budget constraints and, as a result, are below ideal level in relation to their mass testing capacity. Evidences have shown that successful strategies implemented worldwide includes aggressive testing and isolation (18,19), plus the testing capacity is in inverse association to the mortality rate due to COVID-19, which is another indication of the impact of the effective tracing of positive patients (20).

Consistent with data from China, Italy, USA and others (21-23) previous stroke and chronic obstructive pulmonary disease were associated with poor in-hospital survival of COVID-19 positive individuals, together with age. Individuals with previous stroke history can develop severe infection and exhibited neurologic manifestations such as acute cerebrovascular diseases. Chronic obstructive pulmonary disease is characterized by an ongoing immune dysfunction affecting pulmonary and systemic inflammatory mediators. Therefore, these individuals present an increased susceptibility to viral respiratory infections, leading to severe clinical outcomes. Among the symptoms, desaturation and tachypnea were important variables used to predict mortality of COVID-19 patients in comparison to negative patients. The symptoms may vary but it is noticed that fatigue and desaturation can evolve to severe consolidation and pneumonia, acute respiratory distress syndrome (ARDS) and multiple organ dysfunction leading to death (24, 25). Knowledge of these risk factors upon patient admission in cooperation with a rapid diagnosis and appropriate medical management can reduce mortality of COVID-19 patients.

Despite all the efforts, diagnostic reports take days to be avaliable having an excessive impact on hospital’s operating costs when isolating and managing symptomatically patients unnecessarily. Costs during the acute infection are considered to be substantially higher than those for other common infectious diseases, as influenza and pertussis (4-5.5 times higher) (7, 26-28). The direct medical costs are high because a patient with COVID-19 have a greater probability of hospitalization and mortality in comparison to individuals infected with other pathogens. Additionally, positive patients require follow-up care and potentially rehospitalization due to long-lasting damage, with considerable remaining medical costs after the acute infection (26). In our study, patients remained hospitalized for up to 38 days (median 13 days), with 65.5% critically ill needing oxygen supply and intubation. The case fatality rate was 21.8% for confirmed COVID-19. Plus, time to obtain RT-qPCR results ranged from 24.2 to 182.3 hours (median 83.6 hours). During this entire time, hospital needed to decide the safest manner to isolate the suspected patients, with a great chance of having failed and accommodating negative patients with COVID-19 positive patients. In this sense, with transmission rates constantly increasing and hospitals at risk of saturation, a rapid virus detection option is a good strategy to keep COVID-19 positive patients and their contacts properly in isolation receiving adequate health care, and implement immediate control measures to limit the outbreak and the appearance of new epidemic peaks.

Comparable performance of SARS-CoV-2 rapid antigen test was shown in symptomatic individuals who were previously unknown positive patients in a reference hospital in Minas Gerais, Brazil. With RT-qPCR results as the reference standard, the sensitivity of SARS-CoV-2 rapid antigen detection showed to be 69%, being higher on female patients (79%) and during the first days of symptoms onset (≤ 3, 69%; 4-7, 80%). Reports have present an overview of sex-disaggregated data from countries worldwide clearly demonstrating similar numbers of cases in women and men, but an increased case fatality in men (30, 31). This supports the interpretation that a consistent biological phenomenon is operating and is responsible for the largest number of fatal cases in men, independent of country-specific demographics and, most importantly, testing strategies (32-34). Other than this, the demographics and clinical characteristics seems not differ between patients who tested positive and negative for SARS-CoV-2 RNA (35, 36). How long SARS-CoV-2 RNA is present in the upper and lower respiratory tracts and in extrapulmonary specimens remains undetermined (37), and the RT-qPCR sensitivity decreases from ≥ 90% on day 1 to 3 post-symptoms, to nearly 80% on day 6 and < 50% by day 14 (38). More supportive data is needed for a clear comprehension of the kinetics of viral loads based on antigen levels, although a strong correlation between antigen levels and the viral load is found during the clinical course of positive patients, with a similar declining trend after 8 days of symptoms (39). We demonstrated an overall PPV of 97%, that reached 100% when diagnosis were done during the 4th and 15th days post-symptoms, with an accuracy of 90-91% from day 1 to 7 of symptoms. While others found that older age was correlated with higher viral load (18, 39), no difference on diagnosis was found here for groups divided by age.

‘Substantial’ agreement was found between the rapid antigen test and RT-qPCR performed both on the same day (κ = 0.72), an agreement that was maintained for the first seven days of symptoms when diagnosis period were separated. For female patients, an ‘almost perfect’ agreement was found (κ = 0.82) regardless of the number of days and, agreement was reduced to ‘fair’ with a poorest performance of both tests revealing the highest number of discordant results when diagnosis were performed after the 8th day of symptoms. The fact that testing may be initially negative in patients with SARS-CoV-2 infection, especially in those who will later develop overt COVID-19, is not really surprising considering the kinetics of SARS-CoV-2 infection. The incubation period for coronavirus is believed to extend to 14 days, with a median time of 4-5 days from exposure to symptomatic onset (29). It appears that transmission is possible for approximately 8 days after symptoms appear. The period from the first day of detection to virus clearance is usually 12 days in those who became instead symptomatic (40, 41). It is also worth mentioning that virus shedding in some patients may continue for some days after symptom relief (42, 43). Other authors have questioned the low performance of antigen detection as frontline testing but they did not considered the testing period time (4, 39), which we conffirmed here that is imperative for both RNA and antigen detection.

The rapid antigen test reproduced RT-qPCR results on 87.1% of patients and did not showed any inconclusive result, preseting strong colorimetric reaction on positive samples and completely absence of color on the negatives. On the other hand, RT-qPCR presented 11 inconclusive results (7.3% of the cases). Of all these inconclusive cases, 1 was positive for the SARS-CoV-2 rapid antigen test on the first day of symptoms presenting odynophagia and dry cough. Although the other 10 patients were negative for the antigen test, only 2 patients were on the right diagnosis timing on samples collection day (5th day of symptoms). Among others, 2 patients presented chest tomography with bilateral multifocal ground-glass opacities compatible to COVID-19, 4 needed oxygen supply after presenting strong dyspnea and desaturation, and 2 of them evolved to death. Although RT-qPCR testing fills a crucial role in accurately detecting SARS-CoV-2 on a case-by-case basis, it also has inherent problems that limit its utility. Molecular detection of the virus failure are related to the quality of the examined kit as well as to the novel coronavirus’ characteristics, sampling location, sampling volume, transportation, and storage, as well as laboratory test conditions and personnel operation (44). In brief, these typically encompass instrument malfunctioning with inappropriate cycling conditions, use of insufficient or inadequate material, non-specific annealing of PCR to homologous sequences, misinterpretation of expression profiles and so forth (41). It is reported that many suspected cases with typical clinical characteristics of COVID-19 and chest tomography images were not diagnosed by RT-qPCR (45). Results from a technique using primers in different genes can be affected by the variation of viral RNA sequences. False-negative and invalid results may occur by mutations in the primer and probe target regions in the SARS-CoV-2 genome, even when based on the conserved regions of the viral genomes (37). Moreover, RT-qPCR may lead to inconclusive results due to low viral load in very early phase or in late phase of disease, mutation of the virus or other technical difficulties in handling of samples (46).

Together, 17 patients (12.2%) were considered positives on the molecular diagnosis and negatives on the rapid antigen test, while 1 (0.7%) were positive only on the rapid antigen test. Amongst the 17 patients positive only for RT-qPCR, only 6 were diagnosed between days 3 to 8 of symptoms. Seven presented chest tomography compatible to COVID-19 findings, but no lung infiltrate on chest x-ray, and none of the patients presented low saturation or needed ventilation or oxygen supply. Patient that tested positive only for the rapid antigen test were on the second day of symptoms, presented fever, dry cough and dyspnea, and chest tomography with multifocal ground-glass opacities on both sides. As noticed before, a single negative test does not exclude SARS-CoV-2 infection, especially in highly exposed persons, if the test is performed using a nasopharyngeal swab specimen and at the beginning of the infection (37, 44, 45, 46). False negative RT-qPCR would be expected, to an extent difficult to estimate, related to conditions already discussed here and, although the specificity of the RT-qPCR seems to be very high, there may be false positive results due to swab contamination (47).

Limited information was available to describe laboratory exams of the patients requiring hospitalization and it was not possible to analyse a correlation of the exams results to the severity and outcomes. In terms of imaging, the incidence of lung infiltration and bilateral multifocal ground-glass opacities in elderly patients (≥ 60 years) was higher than in young patients, as reported before (48, 49). Amongst RT-qPCR positive patients, 38.8% presented chest tomography alterations in comparison to 16.4% of negative patients with same alterations. These data reinforce that chest imaging is extremely helpful in the diagnosis of the disease and has an irreplaceable role in the early detection as well as monitoring the disease’s clinical course (50, 51).

Management starts with diagnostics and triaging of COVID-19 suspected patients at the unit care. A unique understanding of the tradeoffs inherent in returning to home or an acute hospitalization becomes relevant (52). The medical and health team should be educated regarding disease course, comorbities and symptoms with potential evolution to severe cases, COVID-19 complications. Most importantly, hospital needs to be prepared for a rapid decision about patients isolation. There should be a low threshold for suspicion and confirmation of SARS-CoV-2 infection. To achieve this goal, efforts should be made to conduct testing and management in rapidly accessible areas with a low risk of exposure to restrict exposure to virus and to avoid overwhelming the healthcare system (45, 46, 48). Our observations revealed that the first SARS-CoV-2 antigen test recommended by WHO on the Emergency Use Listing for In vitro Diagnostics (IVDs) Detecting SARS-CoV-2, published at 22^nd^ September, 2020, is a good alternative for immediate diagnosis of COVID-19 at the time of hospitalization of symptomatic and suspected patients. Results suggest that this point of care COVID-19 antigen test has the potential to allow earlier detection for an immediate isolation of positive patients comparable to the time-consuming molecular test, with the aim of reducing in-hospital transmission, allowing proper management of COVID-19 positive individuals.

## Data Availability

All data referred in this paper are available at Oswaldo Cruz Foundation.

## Acknowledgements

We thank Hospital da Baleia care team members for the partnership and for providing an excellent service on the front line in times of COVID-19 and EcoDiagnostica for attending our request for donation of diagnostic kits under evaluation. We also thanks the financial support of Oswaldo Cruz Foundation (to RFQG and scholarships to CC, RA, LC, NC), The Brazilian National Council for Scientific and Technological Development (CNPq) (scholarships to AO, DM, SG), Coordination for the Improvement of Higher Education Personnel (CAPES) (scholarships to NA, JA), The Minas Gerais Research Funding Foundation (FAPEMIG) (scholarship to PF).

